# MiniMORPH: A Morphometry Pipeline for Low-Field MRI in Infants

**DOI:** 10.1101/2025.07.01.25330469

**Authors:** Chiara Casella, Aksel Leknes, Niall J. Bourke, Ayo Zahra, Daniel Cromb, Dora Barnes, Alejandra Martin Segura, Flora Silvester, Vanessa Kyriakopoulou, Daniel Elijah Scheiene, Simone R. Williams, Layla E. Bradford, Joanitta Murungi, Steven C. R. Williams, Sean C. L. Deoni, Victoria Nankabirwa, Kirsten A. Donald, Muriel M. K. Bruchhage, Jonathan O’Muircheartaigh

## Abstract

**Background:** Ultra-low-field (ULF) MRI facilitates neuroimaging access, yet its application in early infancy is constrained by low resolution and contrast, and the limited suitability of existing segmentation tools.

**Objective:** To introduce and validate miniMORPH, an open-source pipeline for automated brain volumetry from 0.064T T2-weighted MRI acquired across infancy and toddlerhood.

**Methods:** ULF scans were acquired from infants aged 2 to 27 months across two cohorts in South Africa and Uganda. Age-specific templates and priors were used to segment major brain tissues and substructures. Validation used two high-field (HF) references: (i) expert manual HF segmentations for key ROIs across ages, and (ii) automated HF segmentations from SuperSynth on paired HF-ULF scans. We quantified (a) between-subject ordering across modalities using Pearson’s correlation (r) and (b) systematic scaling differences using percentage error (PE) and time-corrected percentage error (CPE), stratifying performance by cohort and age. Face validity was also tested via mixed-effects models of age, sex, and birthweight.

**Results:** miniMORPH successfully segmented major brain regions across infancy. In paired HF-ULF comparisons, between-subject ordering was generally preserved across many ROIs, with stronger correspondence in the South African cohort than in the Ugandan cohort at 12 months. Systematic scaling offsets were most evident in CSF-rich or boundary-sensitive compartments, with consistently negative CPE for ventricles and cerebellum. Performance varied with age, showing the greatest variability at 3 months. miniMORPH successfully captured regional age-related growth trajectories. Sex-dependent volumetric differences were widespread but attenuated after intracranial volume correction. Low birthweight infants exhibited reduced regional volumes and altered growth trajectories.

**Conclusion:** miniMORPH enables volumetric analysis of ULF infant MRI and preserves between-subject variation suitable for developmental and group analyses. ROI- and cohort-specific offsets, particularly in CSF-rich regions, may require calibration when absolute volumes are needed. The pipeline is openly available at https://github.com/UNITY-Physics/fw-minimorph.

## 1. Introduction

Infancy is a critical period for brain development, laying the foundation for future cognitive abilities (Bruchhage et al., 2020). MRI provides a powerful tool for visualising this maturation, offering quantitative measures predictive of both current and future cognitive outcomes (Abate et al., 2024). However, access to MRI in low- and middle-income countries (LMICs) remains limited, restricting research capacity and the ability to capture neurodevelopmental variability in these settings (Abate et al., 2024). This gap has major consequences in terms of generalisability of findings: fewer than 3% of available neuroimaging data originates from LMICs, with even fewer studies including children beyond the neonatal period (Rutherford et al., 2022).

Recent advances in portable, ultra-low-field (ULF) MRI (0.064T) offer meaningful potential to expand imaging capabilities in resource-limited or remote environments. These systems are substantially more affordable (∼$200,000-400,000 compared to $1.5-4 million for conventional MRI), require far less power (similar to that of a household appliance), and produce a small fringe field, making them suitable for use in clinical wards or community research centres without the need for extensive magnetic or radiofrequency shielding (Abate et al., 2024). By enabling the acquisition of MRI data in settings where it was previously inaccessible, ULF imaging could significantly advance our understanding of neurodevelopment, particularly in relation to environmental and genetic factors that require large, diverse cohorts to investigate effectively. One such factor is birthweight, a robust marker of early-life health that has been linked to differences in brain volume and structure across childhood (de Kieviet et al., 2012; Walhovd et al., 2012) and may be especially informative in LMICs, where challenges such as malnutrition, infection, and environmental hardship are more prevalent.

Despite its potential, the application of ULF MRI in neonatal and paediatric populations remains limited. To date, only a handful of studies have investigated ULF imaging in these groups (S. C. Deoni et al., 2021; Sabir et al., 2023; Tu et al., 2023), and the suitability of ULF MRI for large-scale research remains an open question. Low anatomical contrast in the infant brain, combined with the lower image resolution and signal-to-noise ratio (SNR) inherent to ULF MRI, present challenges for the application of existing morphometric tools (Donnay et al., 2024). Although established tools such as dHCP (Makropoulos et al., 2018), DRAW-EM (Makropoulos et al., 2014), infant FreeSurfer (Zöllei et al., 2020), and iBEAT (Wang et al., 2023) support age-specific segmentation, and contrast-agnostic frameworks like SuperSynth have improved robustness across different acquisition protocols, these pipelines were primarily developed and validated on high-field (HF), where tissue contrast is higher and anatomical boundaries are better defined. These approaches face substantial limitations and are prone to failure when applied to ULF MRI, raising concerns about reproducibility and interpretability in developmental populations. Dedicated pipelines are therefore needed for realizing the full potential of ULF MRI to support global neurodevelopmental research.

In this work, we present miniMORPH, a template-based pipeline for quantifying brain tissue volumes from 0.064T MRI data acquired across infancy, enabling automatic segmentation of low-resolution T2-weighted (T2-w) images without requiring super-resolution or upsampling. miniMORPH has already shown excellent correspondence between 3T- and 64mT-derived volumes for intracranial volume and key brain regions (including the caudate and putamen) in infants (Ringshaw et al., 2026). Here, we (i) benchmark miniMORPH-derived regional volumes from ULF against two HF reference standards from paired data: expert manual segmentations on the HF images and SuperSynth-derived volumes from the corresponding HF scans; and (ii) assess face validity by testing whether miniMORPH-derived volumes reproduce established biological associations, including age- and sex-related trajectories and birthweight-related differences using mixed-effects modelling.

## 2. Methods

### 2.1. Participants and data acquisition

The data used in this study were acquired at the University of Cape Town (UCT-Khula study) and at Makerere University & Kawempe Referral Hospital (KRH), Kampala (Uganda-PRIMES study). The Makerere University School of Public Health Higher Degrees Research and Ethics Committee (REC reference No: 871) and the Uganda National Council for Science and Technology (UNCST reference No: HS1226ES) gave ethical approval for the Uganda-PRIMES study. The UCT-Khula study received ethical approval from the University of Cape Town Human Ethics Research Committee (HREC; 666/2021). All mothers provided written informed consent for their children.

The Uganda-PRIMES and UCT-Khula cohorts together capture rich developmental sampling across early infancy and toddlerhood. Uganda-PRIMES focuses on the first year of life, with infants recruited as young as two months and follow-up concentrated around 6 and 12 months. The UCT-Khula cohort spans a broader developmental window from 2 to 27 months, with dense representation at 3, 6, 12, 18 and 24 months. Both cohorts include a substantial proportion of children with repeated longitudinal assessments, offering opportunities to study within-child developmental trajectories in addition to cross-sectional age comparisons.

Participants from both cohorts were scanned on the Hyperfine Swoop system (Abate et al., 2024). The protocol included three T2-weighted fast spin echo (FSE) acquisitions in axial, coronal, and sagittal orientations. The axial sequence was acquired with a 112 × 136 × 40 matrix at 1.5 × 1.5 × 5 mm³ resolution (TE/TR = 180/2000 ms, acquisition time = 2:15). The coronal sequence used a 112 × 44 × 124 matrix at 1.5 × 5 × 1.5 mm³ resolution (TE/TR = 220/2000 ms, acquisition time = 2:22). The sagittal sequence was collected with a 36 × 136 × 124 matrix at 5 × 1.5 × 1.5 mm³ resolution (TE/TR = 225/2000 ms, acquisition time = 2:12). Matched HF scans used as reference for validation were collected using cohort-specific HF protocols; details of HF scanner and sequence parameters and the number and age distribution of paired HF-ULF scans are provided in Table 1.

**Table 1:**
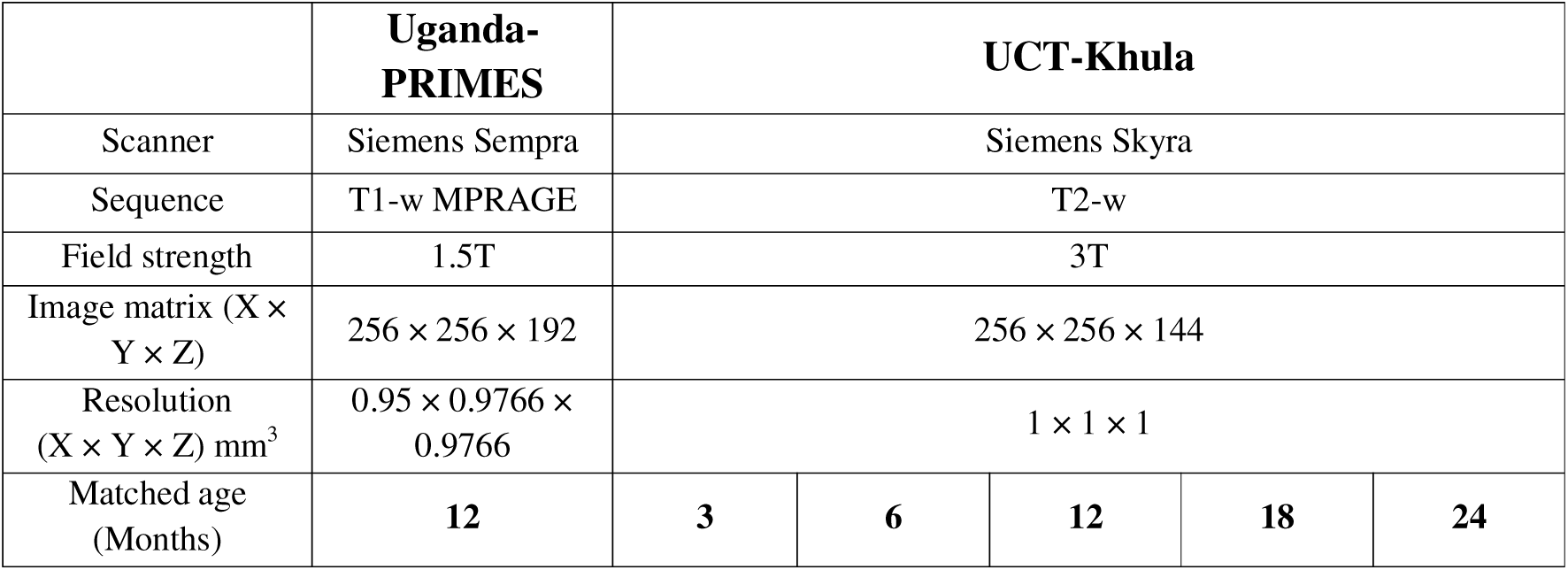
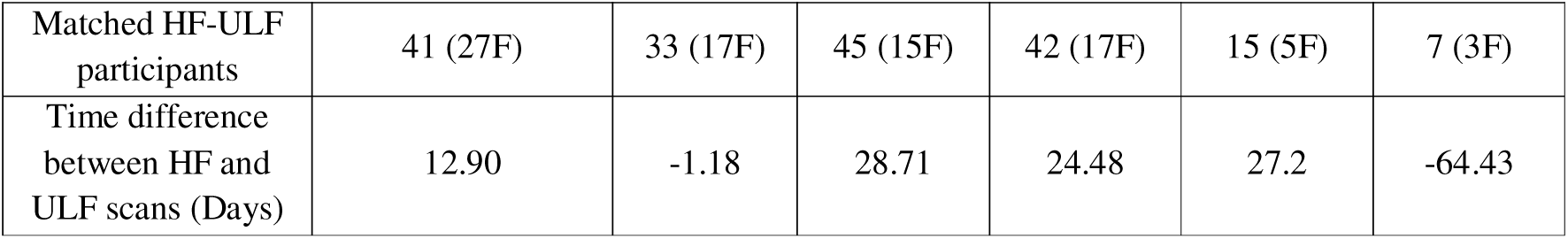
Cohort specifications for Uganda-PRIMES and UCT-Khula with HF scanner specifications and matched participants.

### 2.2. Image processing and segmentation pipeline

miniMORPH uses age-specific templates, tissue priors, and anatomical masks to segment each infant’s ULF T2-w images in native space. The pipeline produces regional volumes for: supratentorial tissue, supratentorial CSF, ventricles, cerebellum, cerebellum CSF, brainstem, brainstem CSF, left and right thalamus, caudate, putamen, globus pallidus, and five corpus callosum segments (posterior, mid-posterior, central, mid-anterior, anterior).

For each scan, a 1.5 mm isotropic image is computed from the three anisotropic T2-w acquisitions and registered to the closest age-specific template. The resulting inverse transformations are then used to warp priors and regional masks into the individual’s native space for segmentation and regional volumetry. Figure 1 summarises the segmentation pipeline.

**Figure 1.**
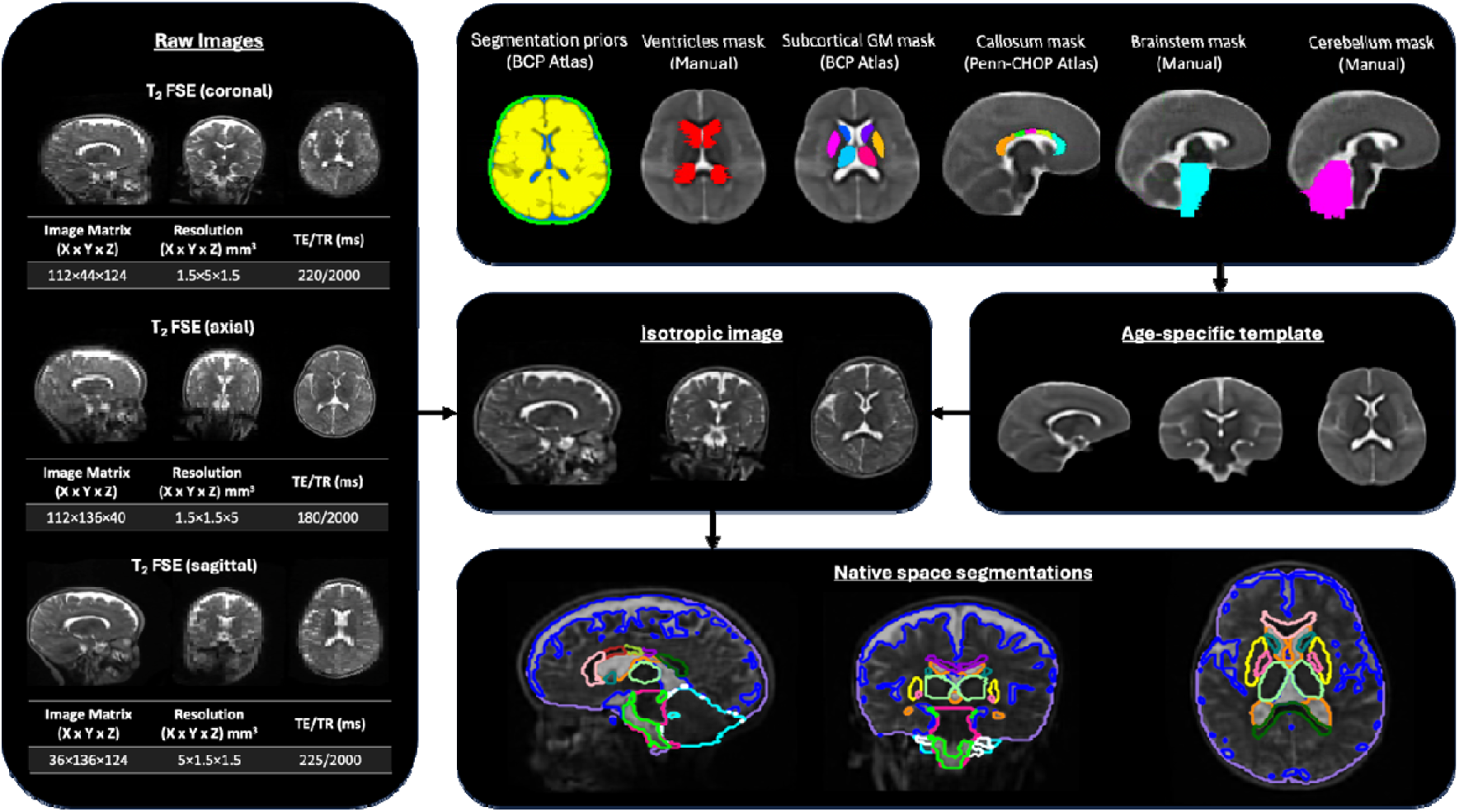
Summary diagram of the segmentation pipeline. The pipeline begins with raw T2-w MRI images acquired in multiple orientations, which are processed to create an isotropic image. Segmentation priors from the BCP atlas (Chen et al., 2022), along with segmentation masks from the BCP and Penn-CHOP atlas (Song et al., 2024), are then aligned to native space through an age-specific template for segmentation.

Below, we describe (i) how isotropic T2-w images were reconstructed and (ii) how the age-specific templates, priors, and masks were generated, followed by (iii) the native-space segmentation and volumetry steps.

#### Computation of T2-w isotropic volumes

Isotropic images to be segmented were produced from the three anisotropic T2-w acquisitions by implementing multi-resolution registration using the Advanced Normalization Tools (ANTs) multivariate template construction pipeline (Avants et al., 2011) as described in previous work (Deoni et al., 2022; Niaz et al., 2022). Here, the low-resolution images are aligned using linear and diffeomorphic registration with symmetric normalization to provide the combined high-resolution isotropic image. Unlike super-resolution, this approach does not synthesise new anatomical detail; it simply aligns and integrates the acquired stacks, reducing the risk of introducing spurious structure.

#### Age-specific template construction

Templates for the 3-, 6-, 12-, 18-, and 24-month age points were constructed using a subset of high-quality, low-motion ULF datasets from the UCT-Khula cohort. Specifically, mri_synthstrip (Hoopes et al., 2022) was first used to brain-extract the isotropic T2-w images. FSL (fslmaths) (Jenkinson et al., 2012) was used to generate edge (gradient) images. Brain-extracted and edge images were then used to create multi-channel templates in ANTs (Avants et al., 2011).

#### Age-specific priors

Tissue and cerebrospinal fluid (CSF) priors were generated by registering T2-w images from the Baby Connectome Project (BCP) atlas (Chen et al., 2022) at 3, 6, 12, 18, and 24 months to the study templates using SynthMorph (Hoffmann et al., 2024). White matter (WM), grey matter (GM), and cerebrospinal fluid (CSF) probability maps were transformed accordingly. A “tissue” prior was created by combining WM and GM maps, and a “skull” prior was generated by dilating the brain mask for improved classification of non-brain regions.

#### Subcortical grey matter masks

To generate subcortical GM masks, age-specific templates were first resampled to 0.5 mm isotropic resolution. The subcortical parcellation maps from the BCP atlas (Chen et al., 2022) were then registered to the 12-month template using the ANTs SyN algorithm (Avants et al., 2011). Thalamus and caudate masks were manually refined on the 12-month template to ensure anatomical accuracy. All resulting masks were subsequently propagated to the remaining age-specific templates using a series of non-linear transformations computed via ANTs.

#### Corpus callosum masks

To obtain callosal parcellations, the callosum mask from the Penn-CHOP Atlas (Song et al., 2024) was first registered to the 12-month template. Manual segmentation of the corpus callosum was then performed on the 12-month template, guided by anatomical landmarks corresponding to the Desikan-Killiany atlas subdivisions (Desikan et al., 2006). The resulting parcellated mask was reviewed for anatomical accuracy and subsequently propagated to the remaining age-specific templates using the same ANTs-based transformation framework applied to the subcortical GM masks.

#### Ventricle, cerebellum, and brainstem masks

Ventricles, cerebellum, and brainstem masks were manually delineated in template space and reviewed by an expert rater. Region-specific strategies were used to optimise segmentation reliability. Specifically, ventricle masks were drawn on each age-specific template and defined to include the bilateral lateral ventricles, third and fourth ventricles, and choroid plexuses. Brainstem and cerebellar masks were initially defined in the 12-month template space and subsequently propagated to the younger and older templates through intermediate age templates (e.g., from 12-month to 24-month via the 18-month template).

#### Individual segmentation and volumetry

For segmentation, T2-w isotropic images were registered to the appropriate age-specific template using ANTs, and the inverse transformations were applied to resample tissue priors into native space. Segmentation into the three primary tissue classes (tissue, CSF and skull) was performed using ANTs Atropos (Avants et al., 2011), with a prior probability weighting of 0.5.

To further refine segmentation, we applied anatomical masks to separate tissue classes spatially. First, we multiplied the CSF tissue class by the ventricle mask to isolate ventricular CSF from extra-ventricular CSF. Next, we used the subcortical GM mask to extract subcortical GM from the overall GM class. Finally, the brainstem and cerebellum masks were applied to both the tissue and CSF posteriors in native space. This procedure allowed us to distinguish supratentorial tissue from brainstem and cerebellar tissue, and to differentiate supratentorial CSF from CSF within the posterior fossa.

### 2.3. Analysis

#### Quality control (QC)

QC of segmentation volumes was performed using outlier analysis against an age-group statistics. Whitin each age bin and each cohort, subjects with greater than ±2 standard deviations in any of the segmented areas were flagged and checked. Participants with poor quality data and segmentations were excluded.

#### HF benchmarking

MiniMORPH was benchmarked against both HF manual segmentations and HF automatic segmentations using SuperSynth.

#### Benchmark 1: Expert manual HF segmentations

To obtain a manual reference, five participants per age group (3, 6, 12, 18 and 24 months) with paired ULF-HF scans were selected from the UCT-Khula dataset. HF scans were manually annotated by a panel of five expert raters. For each participant at each age, two raters independently segmented the ROIs, to enable inter-rater agreement assessment.

We restricted manual annotation to a targeted subset of ROIs to balance anatomical coverage with the practical constraints of manual labelling. The selected ROIs (caudate, putamen, globus pallidus, thalamus, ventricles, and corpus callosum) were chosen to (i) span different tissue compartments (deep GM, WM, and CSF), (ii) prioritise structures that are routinely studied and neurodevelopmentally/clinically salient, and (iii) emphasise regions that are most informative for benchmarking, in particular subcortical areas and CSF spaces where partial-volume effects are more likely. The corpus callosum was assessed as a single structure rather than its five pipeline subsegments to minimise additional manual parcellation time and to avoid introducing extra inter-rater variability.

To quantify uncertainty in manual labelling, the inter-rater relative differences were computed for each region and participant as the absolute relative difference (ARD) in volume between two raters, where *v_1_* and *v_2_* are the volumes estimated from rater 1 and rater 2, respectively. ARD is calculated as follows:

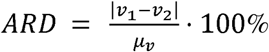

To investigate the difference between HF and ULF segmentations, the percentage error (PE) between HF and ULF was computed as a percentage of the HF reference volume, to enable comparison across regions of different sizes:

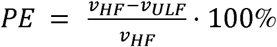

Here, *v_HF_* denotes the HF reference regional volume and *v_ULF_* denotes the corresponding ULF-derived volume. *v_HF_* was taken as the mean of the two raters’ volumes. PE values were then averaged within each age group.

Finally, to capture the extent to which between-subject ordering was preserved across modalities, we computed Pearson’s correlation coefficient (r) between HF manual volumes and ULF miniMORPH volumes for each ROI across the full set of paired scans, pooled across timepoints. Pooling was necessary because only five paired scans were available per age group, which was insufficient to estimate age-stratified correlations reliably.

#### Benchmark 2: SuperSynth-derived HF segmentations

As a second HF reference, we generated automated HF segmentations using SuperSynth, a SynthSeg-based pipeline (Billot et al., 2023; Fischl, 2012), which provides whole-brain parcellations designed to generalise across contrasts and spatial resolutions.

ULF scans were paired with temporally matched HF MRI from the same participants (Uganda-PRIMES: 1.5T; UCT-Khula: 3T). HF scans were acquired within ±90 days of the corresponding ULF scan, with a mean absolute difference ranging from approximately 1 to 29 days across age groups and typically under one month, providing close temporal alignment of the paired acquisitions. Notably, the gap between scans was narrowest at the youngest age points, ensuring minimal developmental change between modalities at these critical early stages. In total, 193 instances of matched HF and ULF scans were available. Age, cohort distribution and mean difference in exact age as well as scanner and sequence information are all shown in Table 1.

Because SuperSynth and miniMORPH use different label sets, output regions were harmonised prior to comparison by aggregating SuperSynth labels into structures matching miniMORPH outputs. The label groupings are reported in Supplementary Table 1. We evaluated SuperSynth-miniMORPH correspondence in the following regions: Cerebellum, ICV (calculated as the sum of supratentorial tissue, supratentorial CSF, cerebellum tissue, cerebellum CSF, brainstem tissue and brainstem CSF), Thalamus, Caudate, Putamen, Globus Pallidus, and Supratentorial Tissue. We excluded inferior ROIs (brainstem and cerebellar/brainstem CSF compartments) from quantitative comparison due to between-scan variability in ULF FOV and coverage.

To assess pipeline performance, the error between paired HF- and ULF-derived regional volumes was measured. PE was calculated for each SuperSynth volume as described for the manual segmentations. As HF and ULF scans were not always acquired on the same day, for this analysis PE was regressed against the time difference between HF and ULF scans (in days) using a linear approximation, yielding a slope *s*. Although early brain growth is non-linear over months to years (Dean et al., 2014), the inter-scan intervals in this study were short (typically less than one month), such that a linear approximation over this restricted time window was considered appropriate. The corrected percentage error (CPE) was then calculated as:

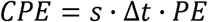

CPE was analysed alongside Pearson’s correlation (r) between matched HF and ULF volumes, which quantifies preservation of between-subject ordering across modalities.

We also quantified cohort differences in pipeline performance by comparing CPE and PE between Uganda-PRIMES and UCT-Khula. Because Uganda-PRIMES contributed matched HF-ULF scans only at 12 months, cohort comparisons were restricted to 12-month-old subjects.

Lastly, age-dependent miniMORPH performance was assessed in the UCT-Khula cohort by computing correlations and CPE separately within each age group (3, 6, 12, 18, and 24 months) across all matched HF-ULF scan pairs.

#### Face validity: effect of age, sex and birthweight

Face validity was first assessed by testing whether miniMORPH-derived volumes recapitulate expected developmental patterns with age and sex in the UCT-Khula cohort using mixed-effects regression models. In this cohort-level modelling, the full set of anatomical regions produced by miniMORPH were included irrespective of whether they were part of the benchmarking subset, because the goal was to interrogate developmental trends in all pipeline-derived measures. Model selection was based on goodness-of-fit, comparing linear versus quadratic age terms for each regional volume using the Akaike Information Criterion (AIC) (Bozdogan, 1987). Volumes were plotted against age in months, with and without correction for ICV.

As an additional face-validity analysis, we also evaluated whether miniMORPH-derived regional volumes were sensitive to the effect of birthweight, categorized as low (Uganda-PRIMES) versus normal (UCT-Khula), on brain tissue volumes. Linear mixed models fitted separately for uncorrected and ICV-corrected regional volumes, with fixed effects for group (low versus normal birthweight), age in months, their interaction, and sex. The UCT-Khula dataset was restricted to participants aged 2-14-months to match sampling and age to the Uganda-PRIMES dataset. To account for multiple comparisons across brain regions, Bonferroni correction was applied to all p-values.

## 3. Results

### 3.1. QC and dataset composition

#Table 2 shows the detailed sample characteristics for each cohort, after QC. Region-wise QC exclusions are reported in Supplementary Table 2. Exclusions varied across regions and ages, with a greater number of excluded segmentations observed at younger ages. Figure 2 illustrates the full parcellation in a representative scan across orthogonal views.

**Figure 2.**
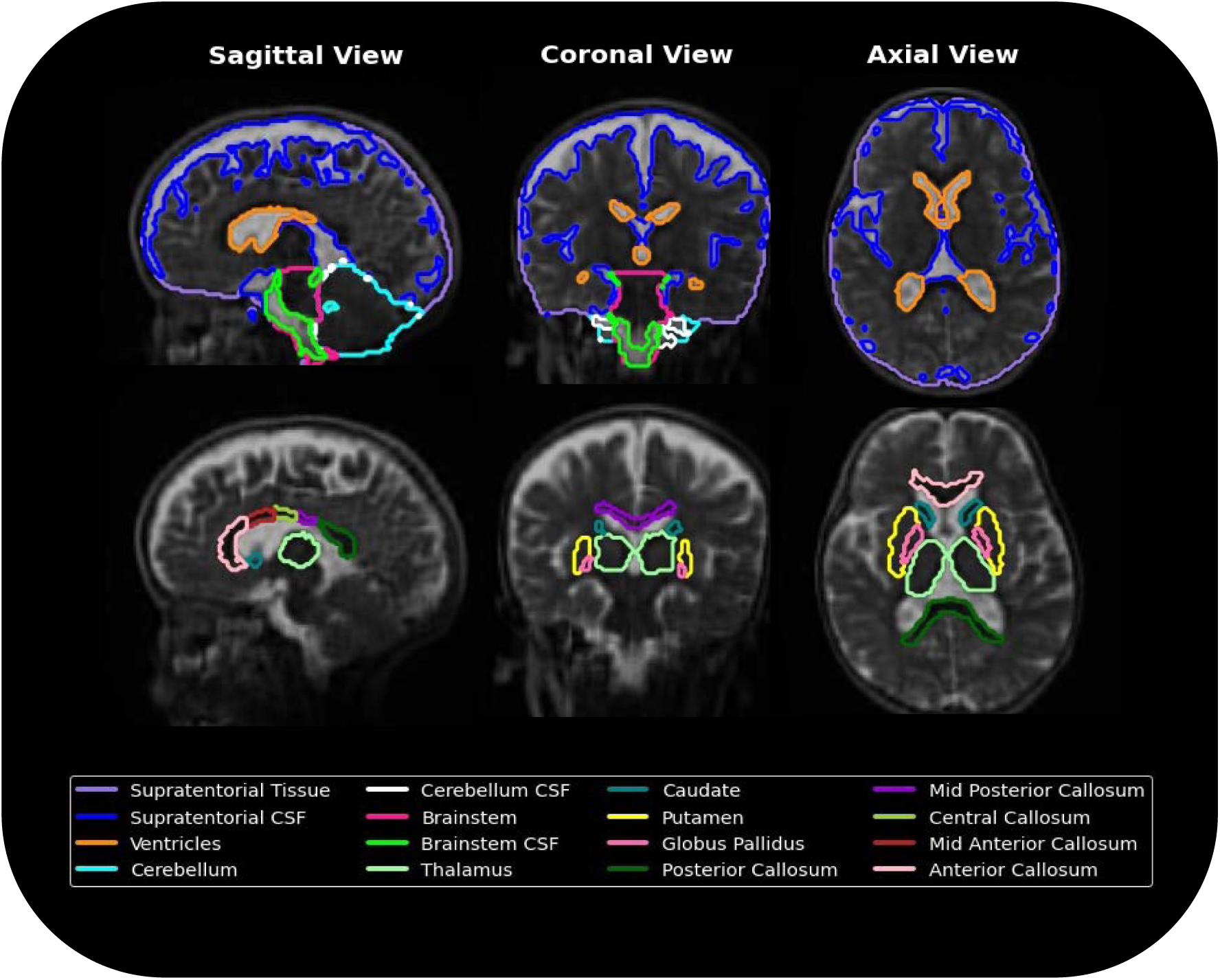
Example segmentation results across sagittal, coronal, axial views. Each view highlights key anatomical structures, demonstrating the detailed parcellation of supratentorial and infratentorial structures critical for volumetric and morphological analysis.

**Table 2.** Sample demographics after QC for each cohort.

| Uganda-PRIMES |  |
| --- | --- |
| Mean Age/Age-range (Months) | 8.2/2-14 |
| Template Age Bins<br>(Scan sessions count) | 3M: 19<br>6M: 52<br>12M: 58 |
| Sex (Count) | 19 Male, 25 Female |
| Subjects with Longitudinal Data | 44 |
| UCT-Khula |  |
| Mean Age /Age-range (Months) | 10.77/2–27 |
| Template Age Bins (Count) | 3M: 105<br>6M: 127<br>12M: 174<br>18M: 87<br>24M: 23 |
| Sex (Count) | 108 Male, 96 Female |
| Subjects with Longitudinal Data | 194 |

### 3.2. HF benchmarking

#### HF benchmarking against manual labels

Table 3A reports the ARD between the two expert raters of each region, averaged across subjects within age group. Inter-rater agreement is generally high for several regions, with particularly low ARD for ventricles (∼3-8%) and putamen (∼2-8%, improving from 12 months onward). In contrast, the corpus callosum shows consistently poor inter-rater agreement across all timepoints (∼27-48% ARD), indicating substantial uncertainty in the manual reference for this structure. Among subcortical ROIs, variability is largest for caudate and globus pallidus, most notably at 6 months (caudate 30.5%; globus pallidus 24.4% ARD), with markedly lower differences from 12 months onwards.

**Table 3:** A) Absolute relative difference (ARD) between two expert raters for each region of interest, averaged across subjects at each timepoint. **B)** Percentage error (PE, %) between manual HF segmentations and miniMORPH ULF segmentations, averaged across subjects at each timepoint. Pearson’s correlation coefficient (r) is reported for each ROI, computed across all timepoints.

| (A) Inter-rater agreement (ARD, %) |  |  |  |  |  |  |
| --- | --- | --- | --- | --- | --- | --- |
| Timepoint | Ventricles | Callosum | Thalamus | Caudate | Putamen | Globus Pallidus |
| 3M | 4.76 | 46.93 | 7.29 | 17.18 | 7.66 | 13.87 |
| 6M | 3.02 | 48.35 | 4.02 | 30.52 | 7.87 | 24.36 |
| 12M | 8.05 | 36.84 | 7.29 | 16.52 | 2.46 | 5.83 |
| 18M | 3.71 | 27.91 | 1.06 | 13.28 | 3.30 | 7.26 |
| 24M | 6.62 | 26.64 | 4.80 | 15.26 | 3.69 | 5.00 |
| (B) Manual HF vs miniMORPH ULF (PE, %), with Pearson $r$ across timepoints | | | | | | |
| | Ventricles<br>$r=0.89$ | Callosum<br>$r=0.72$ | Thalamus<br>$r=0.87$ | Caudate<br>$r=0.87$ | Putamen<br>$r=0.95$ | Globus Pallidus<br>$r=0.87$ |
| 3M | -31.43 | -38.92 | 4.01 | -16.75 | -2.04 | -6.86 |
| 6M | -26.63 | -52.54 | 6.85 | 13.64 | 7.07 | 2.24 |
| 12M | -46.95 | -39.30 | 6.84 | 15.83 | -1.10 | -0.44 |
| 18M | -36.90 | -31.21 | 9.33 | 20.17 | 3.25 | 5.29 |
| 24M | -40.97 | -74.72 | 3.76 | 23.77 | -2.10 | 2.26 |

HF-ULF differences using manual HF as reference are reported in Table 3B. Here, positive values indicate larger HF volumes, whereas negative values indicate larger ULF volumes. Ventricular PE was consistently large and negative (∼−27% to −47%), indicating larger ULF estimates relative to manual HF. Differences for putamen and globus pallidus were small, while thalamus showed a modest positive offset (∼+4% to +9%). Caudate differences changed direction with age (negative at 3 months, positive after), possibly suggesting an age-dependent bias. Callosal PE was large and negative, including an extreme value at 24 months (−74.7%). Given the high manual callosal absolute relative differences (Table 3A), callosal HF-ULF differences should be interpreted cautiously as they combine pipeline error with an unstable manual reference. Overall, unlike the inter-rater results, HF-ULF differences show no clear developmental trend and are broadly consistent across ages.

#### HF benchmarking against SuperSynth

SuperSynth - miniMORPH correspondence across all matched HF-ULF scans for UCT-Khula and Uganda-PRIMES combined is shown in Figure 3. Correlations exceeded 0.8 for all assessed regions, indicating that between-subject variability observed at HF is preserved in ULF-derived volumes. For most ROIs, HF and ULF volumes increased in a similar way across the observed age range. In contrast, the caudate showed a steeper relationship, meaning that the HF-ULF difference becomes larger for participants with larger caudate volumes.

**Figure 3:**
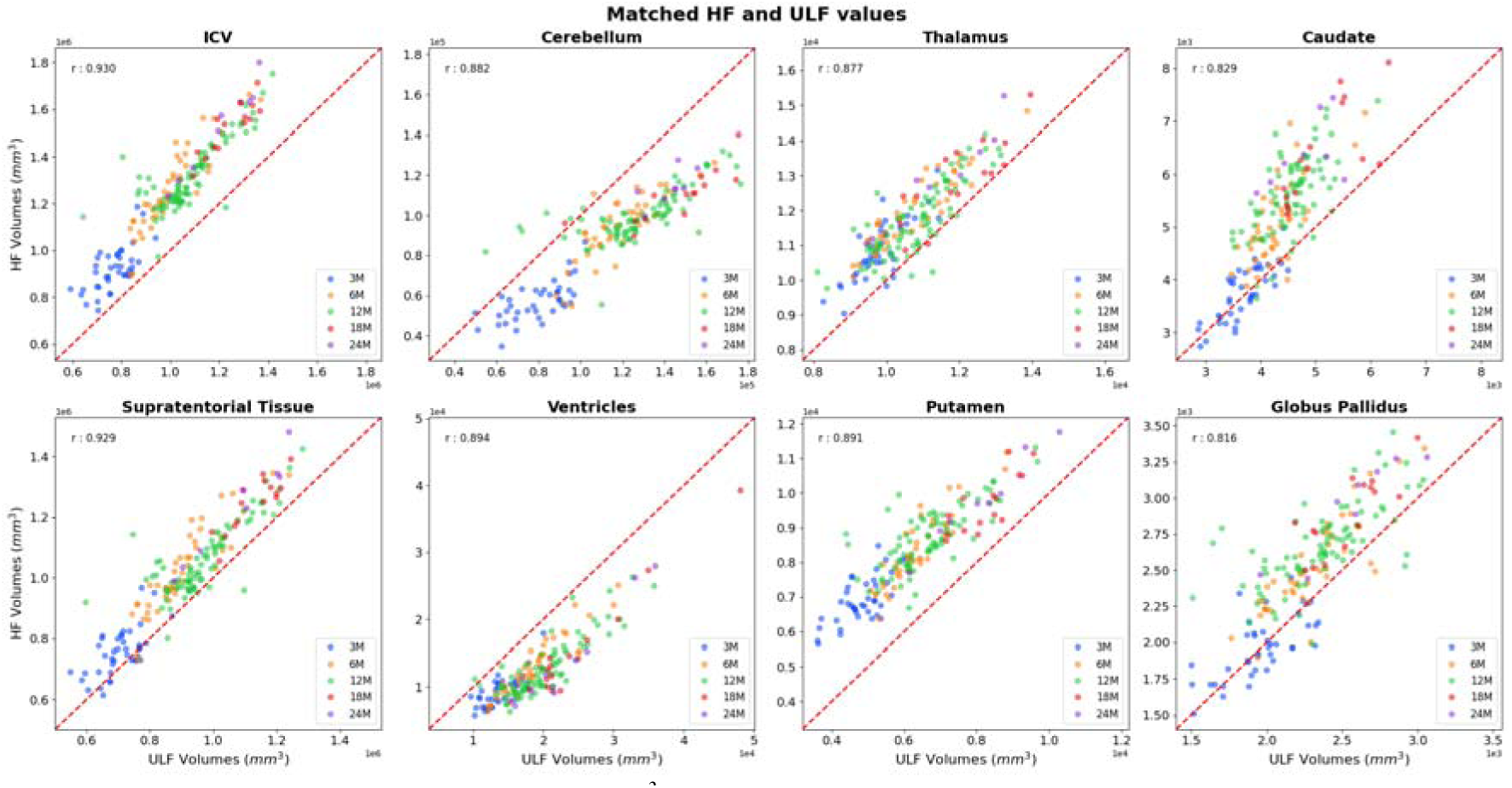
Matched volume estimates in mm^3^ for HF and ULF stratified by region and age. Pearson r is reported for each region across all ages, and the dotted red line represents the identity line.

Looking at CPEs, supratentorial tissue, ICV, thalamus, caudate and globus pallidus all have small positive mean CPEs (∼6-18%), indicating a small bias towards larger HF volumes. Combined with the high correlations reported above, this is consistent with a modest bias toward larger HF volumes while remaining broadly comparable in scale. In contrast, ventricles and cerebellum volumes show negative CPE values, pointing towards larger ULF volume estimates. This is especially true for the ventricles, with mean CPE values below −50% for both cohorts, indicating non-negligible differences in absolute scale between modalities for CSF-rich regions. A complete report of the volumetric discrepancy between ULF-derived segmentations and the HF reference, summarised as MAE (mm³) and MSE (mm⁶) for each region and timepoint, is provided in Supplementary Table 3.

Cohort differences at 12 months are shown in Figure 4 and Table 4. Uganda-PRIMES showed wider CPE distributions (lower precision) across most regions. Cerebellum, putamen and globus pallidus demonstrated large CPE differences between cohorts. Accordingly, the correlations were consistently higher for UCT-Khula than Uganda-PRIMES across all ROIs, indicating stronger linear correspondence between HF and ULF in UCT-Khula at the matched age.

**Figure 4:**
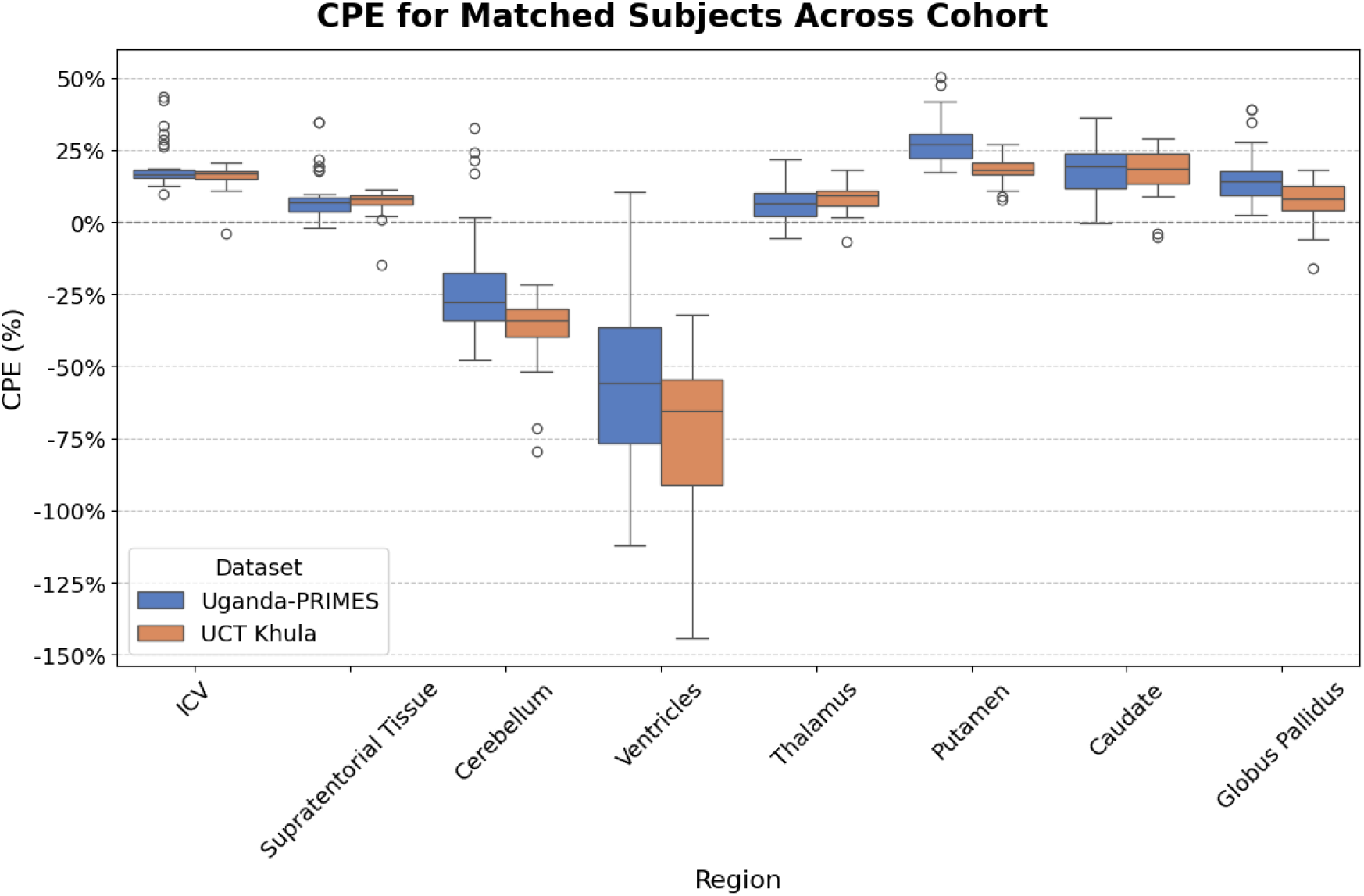
CPE estimates 12 months old subjects stratified by cohort and region. Boxes show interquartile range with median CPE represented as middle line.

**Table 4:** Mean CPE and Pearson’s correlation coefficient for Uganda-PRIMES and UCT-Khula.

|  | Uganda-PRIMES<br>Mean CPE (%) | UCT-Khula<br>CPE (%) | Uganda-PRIMES<br>r | UCT-Khula<br>r |
| --- | --- | --- | --- | --- |
| ICV | 18.733 | 15.980 | 0.365 | 0.914 |
| Supratentorial<br>Tissue | 8.674 | 7.015 | 0.397 | 0.915 |
| Cerebellum | -21.929 | -36.480 | 0.485 | 0.865 |
| Ventricles | -53.138 | -71.778 | 0.830 | 0.942 |
| Thalamus | 6.735 | 8.375 | 0.670 | 0.898 |
| Putamen | 27.678 | 18.232 | 0.518 | 0.908 |
| Caudate | 18.016 | 17.973 | 0.484 | 0.762 |
| Globus Pallidus | 15.158 | 7.436 | 0.587 | 0.756 |

Age-stratified performance in both UCT-Khula and Uganda-PRIMES subjects is show in Figure 5. The spread of CPE is largest at 3 months for all regions except ventricles, indicating that there is an increased random error at 3 months. Most regions have similar performance in terms of correlation and CPE for 6, 12 and 18 months. At 24 months, although a decrease in correlation is observed, the low sample size (N=8) makes performance estimates uncertain.

**Figure 5:**
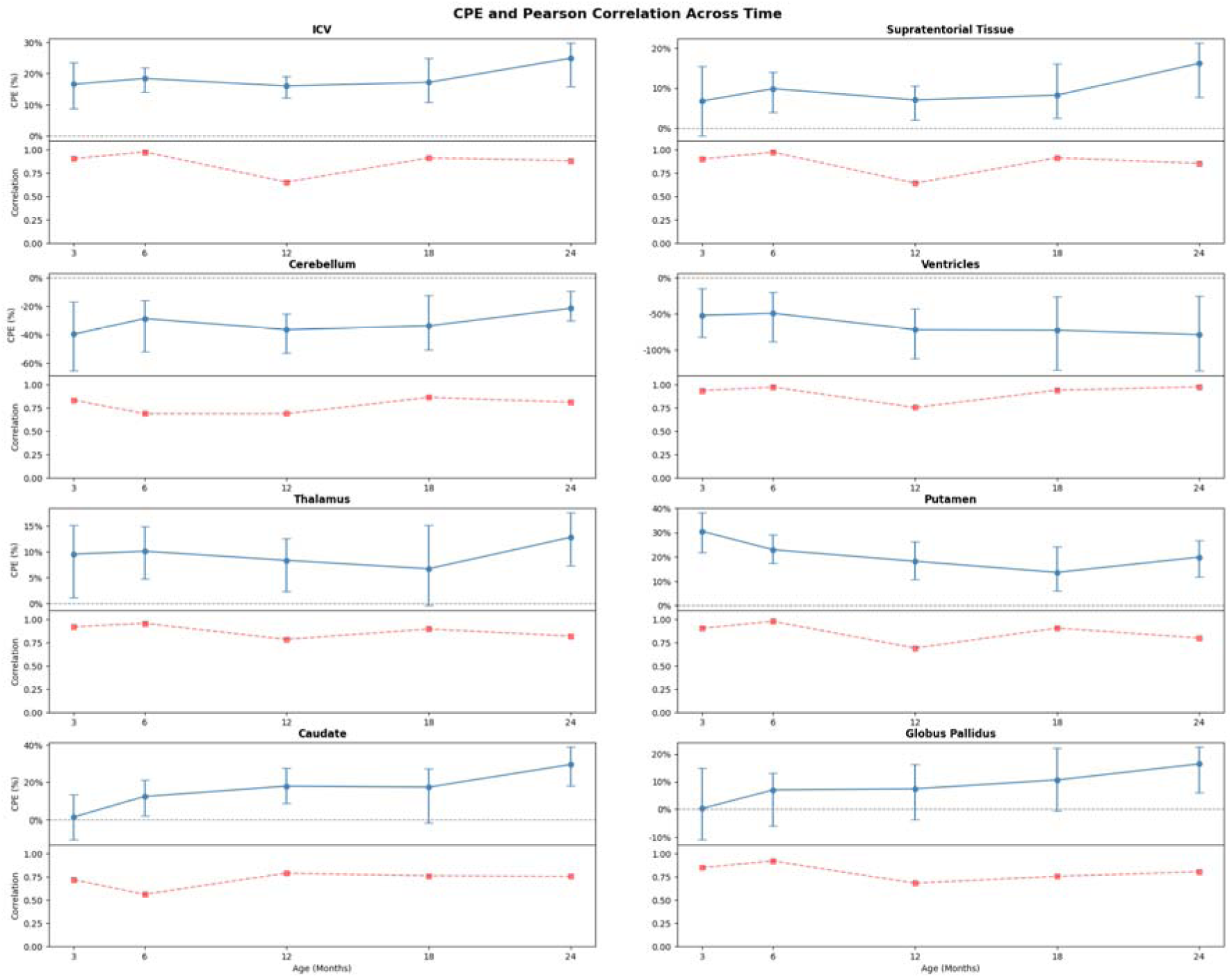
CPE and Pearson correlation coefficient for matched UCT-Khula subjects, stratified by age.

### 3.3. Effect of age, sex and birthweight

#### Age and sex

Age and sex effects for both uncorrected and ICV-corrected volumes are plotted in Figure 6 and detailed in Table 5.

**Figure 6.**
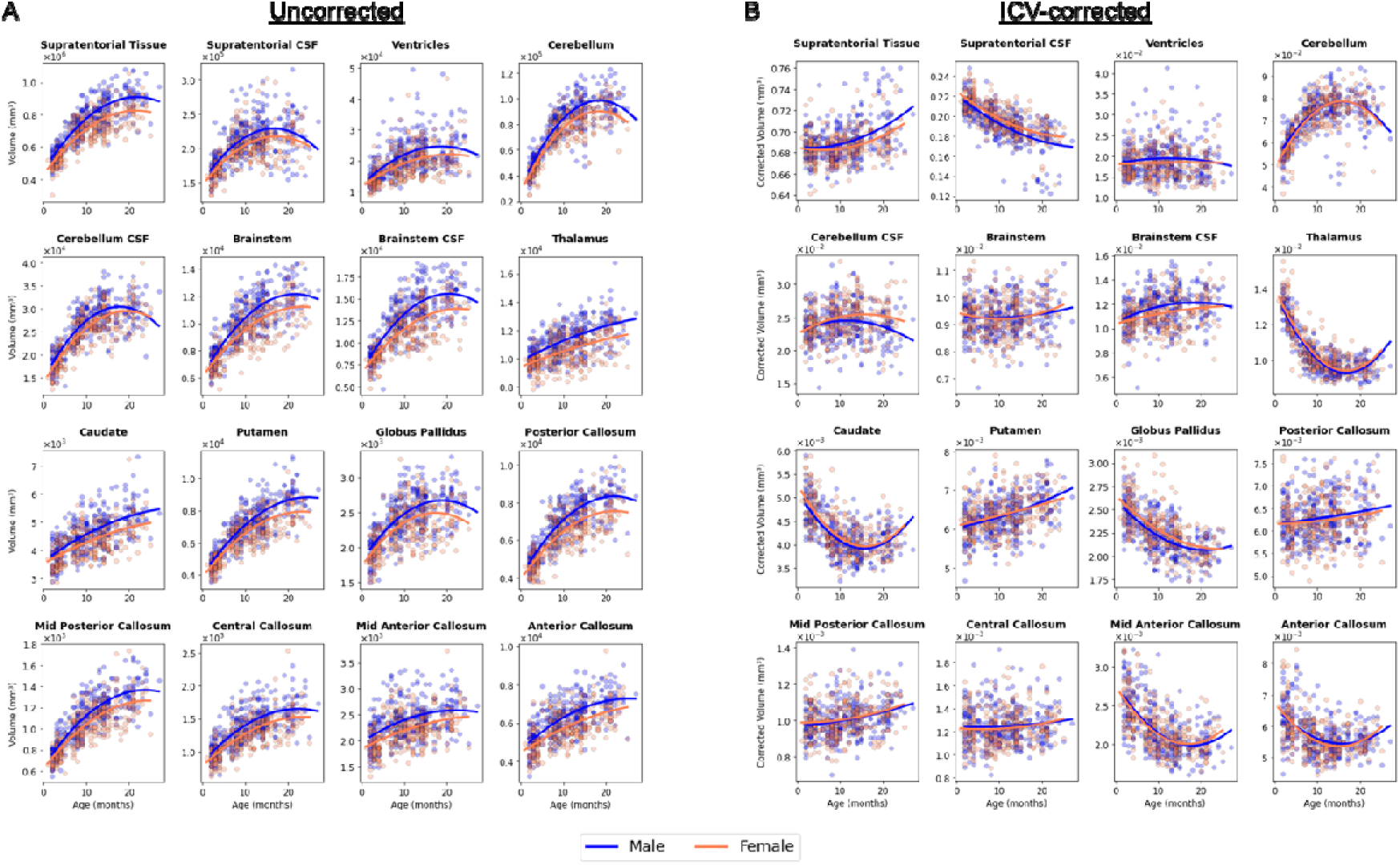
Effect of age and sex on brain volumes. **A)** Absolute volumes. **B)** Volumes corrected for ICV. The datapoints represent individual measurements.

**Table 5.** Summary results of the mixed-effects models assessing the effect of age and sex on regional brain volumes.

| Uncorrected |  |  |  |  |  |  |
| --- | --- | --- | --- | --- | --- | --- |
| Region | Sex Effect | Corrected p (Sex) | Age Effect | Corrected p (Age) | Quadratic Age Effect (p-value) | Corrected p (Age<sup>2</sup>) |
| Supratentorial tissue | 63183.9614 | **2.68E-26** | 39117.4968 | **7.12E-96** | -892.7499 | **3.40E-30** |
| Supratentorial CSF | 10214.0069 | **1.26E-06** | 8791.06053 | **5.63E-45** | -259.3807 | **1.20E-23** |
| Ventricles | 2366.81877 | **3.96E-08** | 1206.84906 | **3.48E-20** | -31.296255 | **2.71E-08** |
| Cerebellum | 7053.71233 | **4.26E-16** | 7231.26777 | **1.88E-163** | -197.21719 | **2.91E-73** |
| Cerebellum CSF | 1309.15432 | **1.46E-06** | 1692.70515 | **4.77E-103** | -45.808409 | **1.61E-45** |
| Brainstem | 782.941131 | **1.50E-15** | 496.848946 | **9.70E-60** | -11.079133 | **3.98E-18** |
| Brainstem CSF | 1472.71826 | **3.09E-21** | 769.7384 | **1.99E-56** | -18.192016 | **3.52E-19** |
| Thalamus | 728.780705 | **1.47E-19** | 107.058132 | **2.16E-60** |  | 1 |
| Caudate | 282.325646 | **6.08E-11** | 100.165797 | **2.50E-13** | -1.3969815 | 0.13967485 |
| Putamen | 520.590705 | **2.52E-14** | 371.001492 | **2.53E-69** | -7.6235112 | **6.79E-18** |
| **Globus pallidus** | 130.705346 | **6.51E-09** | 91.1647054 | **4.61E-41** | -2.5102742 | **6.76E-19** |
| **Posterior callosum** | 589.524984 | **1.47E-19** | 364.310847 | **1.53E-68** | -8.2974108 | **1.21E-21** |
| **Mid-posterior callosum** | 76.7037258 | **6.56E-12** | 57.3489501 | **3.81E-63** | -1.1905914 | **1.25E-16** |
| **Central callosum** | 117.452057 | **1.72E-11** | 68.3637674 | **2.13E-37** | -1.4867587 | **7.99E-11** |
| **Mid-anterior callosum** | 182.547097 | **3.02E-11** | 49.9692895 | **2.83E-08** | -1.0168306 | **0.04515241** |
| **Anterior callosum** | 493.51014 | **5.73E-14** | 182.569712 | **1.12E-18** | -3.2440844 | **0.00115071** |
| **ICV-corrected** |  |  |  |  |  |  |
| **Supratentorial tissue** | 0.00443259 | **0.01142901** | -0.0005605 | 1 | 6.70E-05 | **0.00151859** |
| **Supratentorial CSF** | -0.0053064 | **1.35E-05** | -0.0034477 | **3.66E-22** | 5.97E-05 | **0.00039885** |
| **Ventricles** | 0.00076109 | 0.3123177 | 1.39E-05 | 1 |  | 1 |
| **Cerebellum** | 0.00064017 | 1 | 0.00364034 | **9.26E-90** | -0.000115 | **6.23E-54** |
| **Cerebellum CSF** | -0.0006557 | 0.055598 | 0.00033732 | **3.90E-05** | -1.27E-05 | **0.0002275** |
| **Brainstem** | -1.84E-05 | 1 | 7.64E-06 | 1 |  | 1 |
| **Brainstem CSF** | 0.00047016 | **0.00062178** | 6.18E-05 | **8.47E-10** |  | 1 |
| **Thalamus** | -0.0001369 | 0.19132756 | -0.0005496 | **3.19E-215** | 1.64E-05 | **2.37E-115** |
| **Caudate** | -7.34E-05 | 0.13909017 | -0.0001628 | **1.91E-72** | 5.18E-06 | **3.40E-44** |
| **Putamen** | -2.48E-05 | 1 | 3.22E-05 | **8.96E-24** |  | 1 |
| **Globus pallidus** | -5.50E-05 | **0.00408543** | -4.97E-05 | **7.32E-24** | 1.11E-06 | **2.53E-07** |
| **Posterior callosum** | 6.15E-05 | 1 | 1.30E-05 | **0.001245** |  | 1 |
| **Mid-posterior callosum** | -7.32E-06 | 1 | 4.03E-06 | **3.13E-09** |  | 1 |
| **Central callosum** | 1.61E-05 | 1 | 2.24E-06 | 0.5562203 |  | 1 |
| **Mid-anterior callosum** | 7.64E-06 | 1 | -8.91E-05 | **4.26E-42** | 2.55E-06 | **9.94E-21** |
| **Anterior callosum** | 3.60E-05 | 1 | -0.0001639 | **4.19E-29** | 5.29E-06 | **2.14E-18** |

Nearly all regions exhibit significant positive linear effects of age. Quadratic age terms are significantly negative across most regions, especially in the cerebellum, CSF spaces, and callosal subregions, suggesting nonlinear growth curves with early acceleration and later deceleration. Notably, the thalamus and caudate fit a linear model with a strong age effect and no significant quadratic age term - consistent with more stable linear growth.

After correcting for ICV, age effects in supratentorial tissue become non-significant (p=0.18), likely due to the supratentorial region constituting a large proportion of total ICV. The quadratic term remains marginally significant. On the other hand, the cerebellum retains highly significant age and age² effects post-ICV correction, showing continued region-specific maturation beyond global size scaling. Likewise, the thalamus, globus pallidus and caudate also retain strong age effects after ICV correction, suggesting these structures undergo meaningful development in volume even when overall brain size is accounted for.

Although highly significant in uncorrected models, most callosal regions show reduced or non-significant age effects after correction. The mid-anterior and anterior callosum remain significant for age and age², consistent with regionally specific shape and thickness changes observed in neonatal callosal development.

While sex has robust effects throughout most of the brain for uncorrected volumes — most prominently in supratentorial tissue, cerebellum, and posterior corpus callosum — with males typically having larger regional volumes, as expected, many sex differences are attenuated or eliminated after adjusting for ICV, indicating that most regional differences are proportional to overall brain size.

#### Birthweight

Birthweight effects for both uncorrected and ICV-corrected volumes are plotted in Figure 7 and detailed in Table 6.

**Figure 7.**
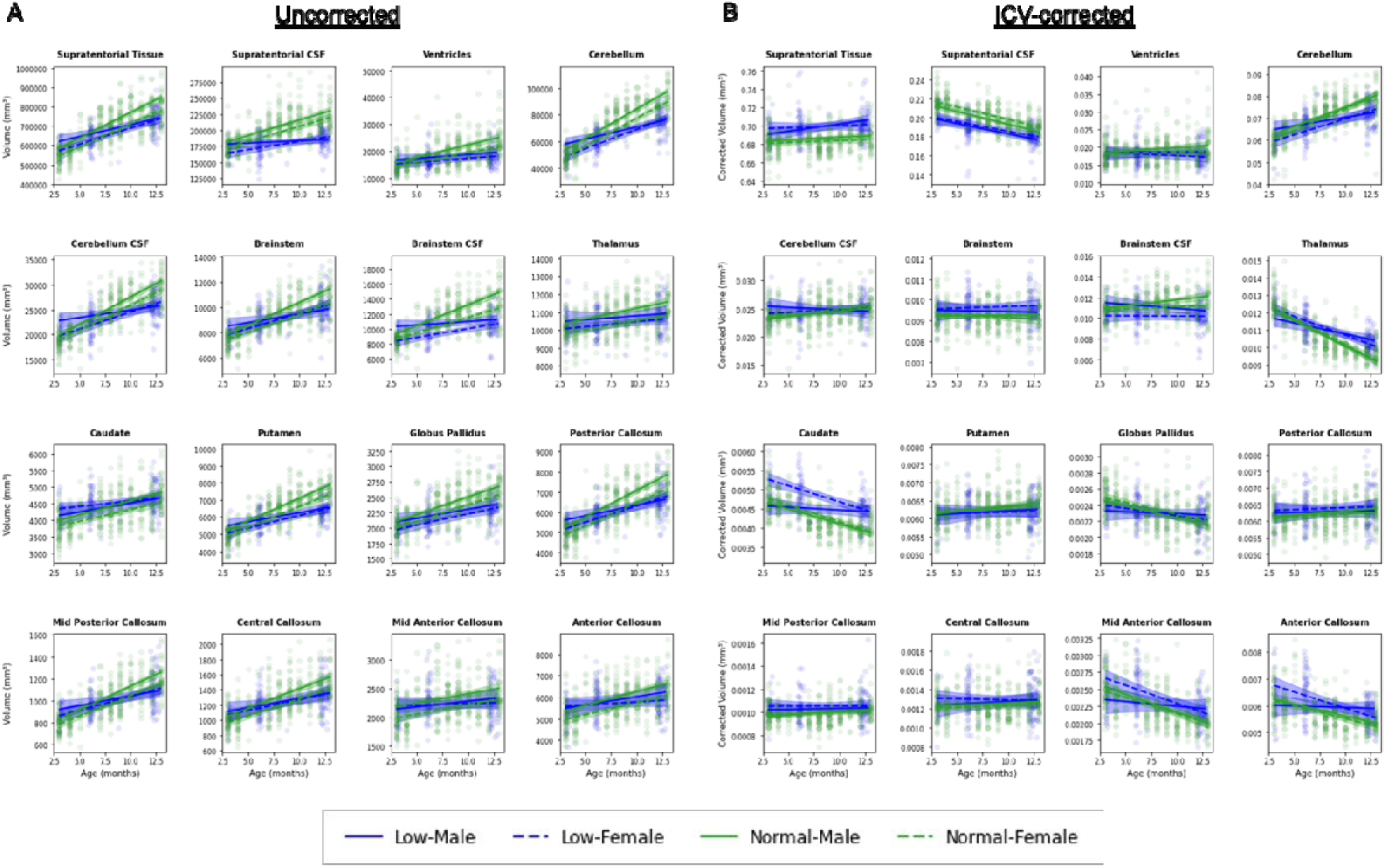
Effect of birthweight on brain volumes. **A)** Absolute volumes (in mm³). **B)** Volumes corrected for ICV. The datapoints represent individual measurements. Each line represents the group means. Shaded areas indicate the 95% confidence interval for each group.

**Table 6.**
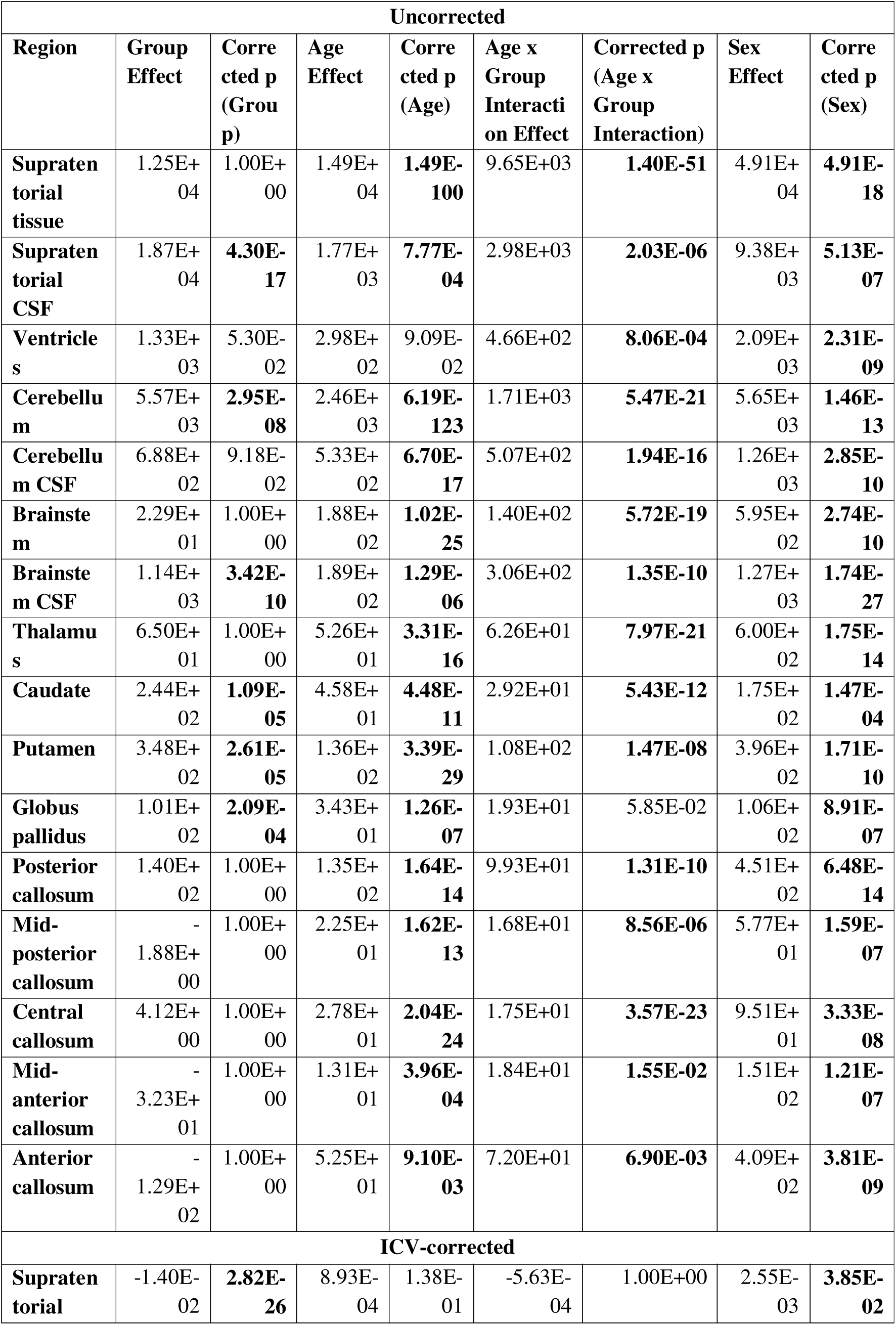
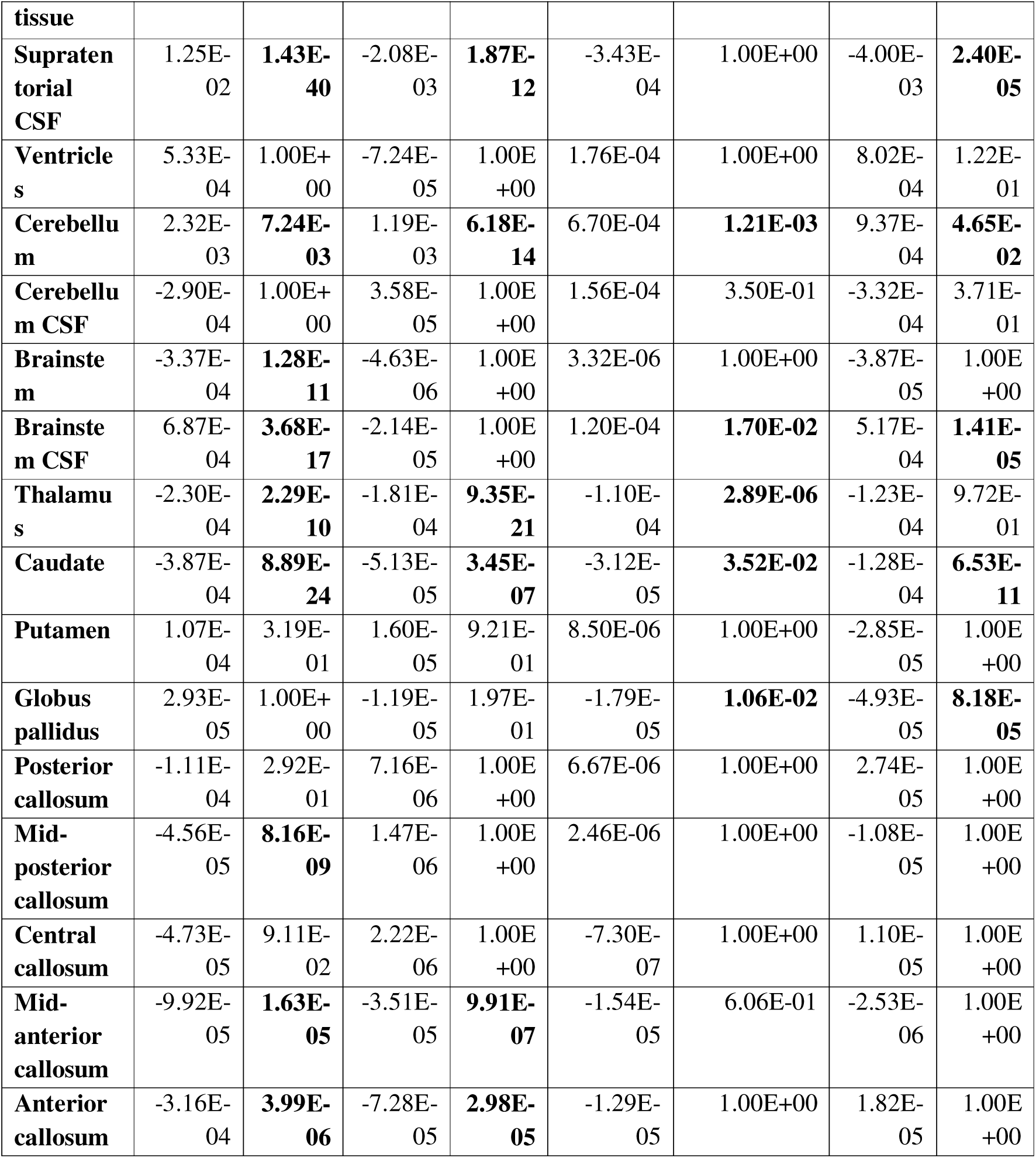
Summary results of the mixed-effects models assessing the effect of birthweight on brain volumes.

Significant effects of birthweight group were observed in multiple brain regions. In uncorrected models, children born with low birthweight exhibited increased CSF volumes and reduced tissue volumes in regions including the supratentorial compartment, cerebellum, caudate, and putamen (*p* < 0.001). These group differences remained statistically significant after ICV correction in selected regions, notably the cerebellum and caudate, indicating localised volumetric alterations not fully attributable to global brain size.

Age-by-group interaction effects were detected in regions such as the cerebellum, thalamus, and corpus callosum, suggesting that developmental trajectories differ by birthweight status. In these regions, volumes in the low birthweight group showed divergent age-related patterns, consistent with atypical or delayed postnatal growth. Age-related increases in brain volume were widespread across the cohort, but birthweight-specific effects and interactions were prominent in both subcortical and callosal structures.

## 4. Discussion

In this work, we introduce miniMORPH, a scalable, fully automated pipeline for extracting brain morphometry from ULF MRI data acquired in infancy. We evaluated the pipeline’s performance using two complementary approaches: (i) benchmarking against HF-derived references, quantifying both preservation of between-subject variation and differences in scaling; and (ii) face-validity analyses, testing whether miniMORPH-derived volumes follow expected age-, sex-, and risk-related developmental patterns.

### 4.1. Benchmarking against manual HF references

Benchmarking against expert manual HF segmentations provided an interpretable reference while also enabling us to quantify uncertainty in the HF benchmark via inter-rater agreement. Agreement was high for ventricles and putamen, whereas the corpus callosum showed consistently poor inter-rater agreement (ARD ∼27-48% across ages), implying substantial instability in the manual reference for callosal boundaries in this dataset. HF-ULF PE patterns were also ROI-dependent: ventricular PE was consistently large and negative (ULF larger than HF), the thalamus showed a modest positive offset (HF larger than ULF), and putamen and globus pallidus had relatively small mean differences. Correlation with manual HF volumes remained high for most ROIs when pooling across ages (for example, putamen r = 0.95; ventricles r = 0.89; thalamus/caudate/globus pallidus r = 0.87), indicating that miniMORPH largely preserves between-subject ordering even when absolute scaling differs.

Crucially, benchmarking metrics should be interpreted in light of the reliability of the HF reference. In regions with low inter-rater variability (for example, the ventricles in our manual benchmark), HF-ULF discrepancies are more likely to reflect method-specific differences. In regions with high inter-rater variability (for example, the corpus callosum), apparent HF-ULF disagreement is harder to interpret because reference uncertainty and pipeline error are partially conflated.

### 4.2. Benchmarking against SuperSynth-derived HF references

Benchmarking against SuperSynth-derived HF segmentations indicated cohort-dependent cross-modality correspondence (Table 4). In UCT-Khula, HF-ULF correlations were high across all assessed ROIs (r = 0.756-0.942), consistent with strong preservation of between-subject differences. In contrast, correlations were more variable in Uganda-PRIMES at 12 months: associations in the ventricles remained high (r = 0.830), were moderate in the thalamus (r = 0.670), whereas other regions showed low-to-moderate correspondence (r = 0.365-0.587; e.g., ICV, supratentorial tissue, cerebellum, caudate, putamen). This pattern suggests that while relative ordering is well preserved in UCT-Khula, cross-modality consistency is reduced in Uganda-PRIMES for multiple ROIs. Consistent with the cohort-specific correlation patterns, cohort stratification also revealed systematic differences in CPE that were structure-specific: Compared to Uganda-PRIMES, UCT-Khula showed smaller positive biases for putamen and globus pallidus (closer to zero), but a larger negative bias for cerebellum.

Several sources of heterogeneity may underlie these cohort effects, most notably differences in the HF reference acquisitions. UCT-Khula participants were scanned at 3T with T2-w contrast, whereas Uganda-PRIMES participants were scanned at 1.5T with a T1-w sequence. Lower field strength is expected to reduce SNR, potentially decreasing the reliability of HF-derived benchmarks, while contrast differences can shift boundary placement, especially near CSF interfaces. In addition, both cohorts were registered to templates constructed from UCT-Khula ULF data. Although the same ULF sequence and scanner were used for both cohorts, demographic, technical, or procedural differences might have propagated into the template space and increased registration error when applied to Uganda-PRIMES. Overall, these cohort-dependent results mirror previous multi-site infant segmentation benchmarks reporting notable drops in Dice when models are tested out of site (Sun et al., 2021).

Across cohorts, mean CPE deviation was most pronounced in the ventricles. Two factors likely contribute to this result. First, SuperSynth and miniMORPH differ in both modelling approach and label definitions: while miniMORPH uses infant-specific labels, the current version of SuperSynth is trained on labels from adults and children, but not infants. This difference might generate a systematic shift by dilating or eroding segmented areas. Second, the ventricular bias is consistent with the acquisition sequence used for the ULF scans: ULF data were acquired with T2-w contrast, where CSF is hyperintense. Combined with lower SNR and reduced spatial resolution, this increases partial-volume effects at CSF-tissue interfaces and may lead to an overestimation of ventricular volume, especially in participants with small ventricles. Such apparent ventricular enlargement would result in a negative CPE, which aligns with our observed findings. Importantly, this explanation may also help interpret patterns in structures bordering CSF, including the caudate and cerebellum. If ventricular boundaries are systematically shifted at ULF, neighbouring ROIs can inherit correlated boundary ambiguities, which can inflate apparent discrepancies even when overall between-subject ordering remains preserved.

miniMORPH performance varied with developmental stage. CPE variance was highest at 3 months across regions, indicating greater uncertainty early in infancy. This likely reflects a combination of neurodevelopmental and technical factors. Between approximately 3 and 9 months, ongoing and spatially heterogeneous myelination reduces definition of the grey-white boundary, lowering tissue contrast and increasing segmentation ambiguity. The reduced contrast at this stage challenges both convolutional segmentation approaches, such as those used in SuperSynth, and registration-based pipelines like miniMORPH, and are not unique to ULF imaging (Li et al., 2019; Sun et al., 2021). Additionally, HF and ULF segmentations are influenced by different imaging properties (contrast mechanisms, SNR, resolution), so their error sources are not expected to be strongly correlated. This uncorrelated noise propagates into the CPE and the normalisation by HF volume further inflates relative variability for small structures and at younger ages. Because CPE is scaled by the HF volume, smaller ROIs provide smaller denominators, so the same absolute discrepancy translates into a larger percentage difference, particularly in early infancy when regional volumes are smallest.

### 4.3. Summary and practical implications of benchmarking results

Overall, benchmarking against HF-derived references indicates that miniMORPH preserves between-subject variation across many ROIs, supporting its use for analyses driven by relative differences (for example, developmental trajectories and group comparisons), even when absolute volumes differ between field strengths. This conclusion is supported by both the targeted manual benchmark and the broader SuperSynth benchmark, but the SuperSynth results also show that cross-modality correspondence is cohort-dependent. Specifically, miniMORPH showed the strongest correspondence in the contrast-matched 3T T2-w benchmark (UCT-Khula), whereas comparisons against the 1.5T T1-w HF reference (Uganda-PRIMES) were more variable, consistent with greater reference heterogeneity and contrast-driven boundary differences.

Systematic ROI- and cohort-specific offsets (captured by CPE) further indicate that agreement in absolute volume is region- and cohort-dependent. For example, supratentorial tissue showed high correspondence and low bias, whereas ventricles showed strong correspondence but a consistent negative bias, plausibly driven by modality-specific contrast and partial-volume effects at CSF boundaries. Where absolute volumes are required, these offsets can be modelled using ROI- and cohort-specific calibration (for example, linear mapping), ideally estimated on training data and validated out-of-sample to assess generalisability across ages and sites.

### 4.4. Face validity: developmental, sex, and risk effects

Having established miniMORPH’s performance and its cohort- and ROI-dependent limitations, we next assessed face validity by testing whether miniMORPH-derived volumes recapitulate established developmental patterns. The pipeline successfully captured significant age-related volumetric changes in both supratentorial and infratentorial regions, as well as subcortical nuclei such as the caudate and putamen. Notably, these effects remained significant for many structures after ICV correction, suggesting the pipeline is sensitive not only to global brain growth but also to region-specific maturational changes. The inclusion of non-linear age terms further highlighted the pipeline’s capacity to resolve developmental acceleration and deceleration phases typical of early infancy (Dean et al., 2014). Together, these results demonstrate that miniMORPH achieves biologically meaningful segmentation of ULF data, enabling robust volumetric analyses during a critical window of brain development.

Sex-related differences were also evident across widespread brain regions in uncorrected volumes, with males generally exhibiting larger volumes in structures such as supratentorial tissue, the cerebellum, and posterior corpus callosum (Gilmore et al., 2007; Knickmeyer et al., 2014). After correction for ICV, most of these differences were substantially attenuated, indicating that much of the observed variance reflects overall head size rather than regional specialisation. Residual effects that remained significant after normalisation were modest and were largely restricted to CSF spaces and selected subcortical structures. This suggests that global scaling effects drive much of the apparent sexual dimorphism in neonatal brain volumes, with few brain region specific differences persisting after normalisation. Overall, these findings are consistent with previous evidence that volume-normalised comparisons reveal few significant sex differences in early life (Holland et al., 2014; Knickmeyer et al., 2014).

As an additional sensitivity analysis, we examined whether miniMORPH-derived volumes detect group-level differences in a low versus normal birthweight comparison. Significant effects of birthweight group were observed across multiple regions. In uncorrected models, children born with low birthweight exhibited increased CSF volumes and reduced tissue volumes in regions including the supratentorial compartment, cerebellum, caudate, and putamen. After ICV correction, birthweight differences persisted in selected regions, notably cerebellum and caudate, suggesting that group effects were not fully explained by global head size. Age-by-group interaction effects were also identified in structures such as the cerebellum, thalamus, and corpus callosum, suggesting divergent developmental trajectories associated with birthweight. These results support sensitivity of the pipeline to clinically relevant group variation and underscore the potential of miniMORPH for early identification of infants at risk for adverse outcomes, particularly in settings where HF imaging and long-term follow-up may not be feasible. Nevertheless, it should be noted that birthweight was confounded with acquisition site in this proof-of-concept analysis. Future work should aim to replicate and refine these findings in harmonised, multi-site cohorts.

### 4.5. Conclusions

In summary, miniMORPH provides a scalable automated pipeline for extracting infant brain morphometry from ULF MRI without requiring super-resolution or upsampling, enabling automated segmentation and volumetry in settings where conventional HF neuroimaging is infeasible. Despite the inherent limitations of ULF imaging, namely lower resolution, SNR and contrast, miniMORPH outputs a comprehensive set of regional measures spanning major tissue and CSF compartments, deep grey matter nuclei, and corpus callosum subregions, and yields developmental trajectories that align with established early-life patterns.

Across both benchmarking approaches, miniMORPH preserves between-subject variation most consistently when evaluated against contrast-matched HF reference data (3T T2-w), supporting its use for detecting volumetric differences and developmental change (for example, age-related growth and group effects) even when absolute scaling differs between modalities. Absolute volume comparability remains ROI- and cohort-dependent, but systematic offsets can be addressed via ROI- and cohort-specific calibration models validated out-of-sample.

Taken together, these findings establish miniMORPH as a practical tool for large-scale, early-life neurodevelopmental studies using ULF imaging. The pipeline bridges a critical methodological gap by allowing robust volumetric analysis in contexts where traditional HF neuroimaging is infeasible, thereby advancing efforts toward more equitable and accessible pediatric brain health monitoring. The pipeline is openly available at https://github.com/UNITY-Physics/fw-minimorph.

## Data Availability

The pipeline presented here is openly available at: https://github.com/UNITY-Physics/fw-minimorph. Other code excerpts, information regarding the analysis, or intermediary results can be made available upon request to. The UCT-Khula data utilised for part of the analysis have been deposited in ZivaHub Open, a public repository. This information can be found at the following DOI: https://zivahub.uct.ac.za/articles/dataset/Khula_South_Africa_Dataset_Iron_Deficiency_Anaemia_with_Adjustment_for_Inflammation/28706561

## Acknowledgements

The authors thank the caregivers, families, and infants in South Africa and Uganda who made this work possible. We would also like to acknowledge the Khula South Africa and Uganda data collection and data management team for the many hours that went into acquiring the data used in manuscript. SW would like to thank the National Institute for Health and Care Research (NIHR) Maudsley Biomedical Research Centre (BRC) for their ongoing support of KCL neuroimaging facilities.

## 8. Supplementary Information

**Supplementary Table 1.**
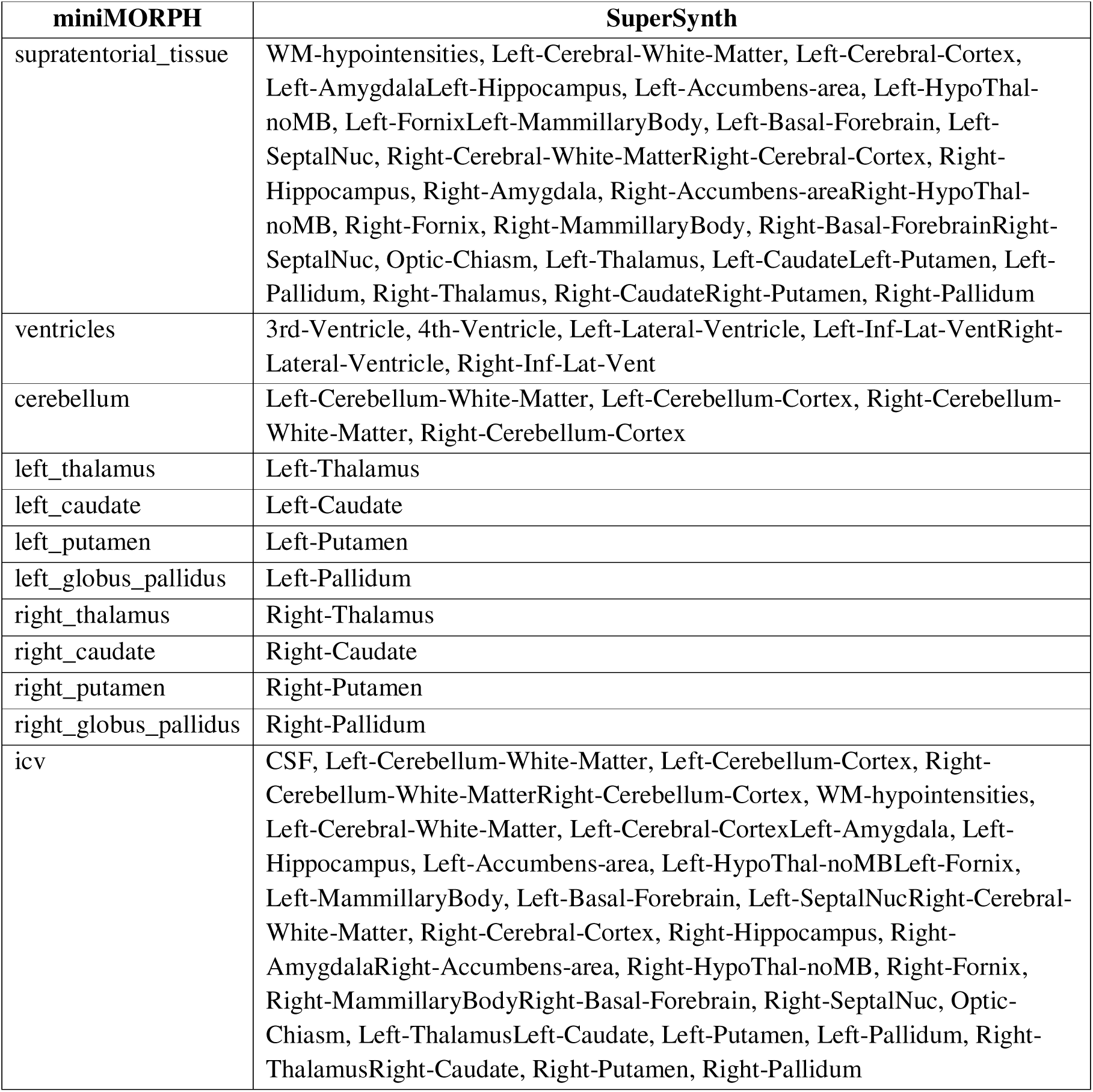
ROI harmonisation between miniMORPH and SuperSynth.

| miniMORPH | SuperSynth |
| --- | --- |
| supratentorial_tissue | WM-hypointensities, Left-Cerebral-White-Matter, Left-Cerebral-Cortex, Left-AmygdalaLeft-Hippocampus, Left-Accumbens-area, Left-HypoThal-noMB, Left-FornixLeft-MammillaryBody, Left-Basal-Forebrain, Left-SeptalNuc, Right-Cerebral-White-MatterRight-Cerebral-Cortex, Right-Hippocampus, Right-Amygdala, Right-Accumbens-areaRight-HypoThal-noMB, Right-Fornix, Right-MammillaryBody, Right-Basal-ForebrainRight-SeptalNuc, Optic-Chiasm, Left-Thalamus, Left-CaudateLeft-Putamen, Left-Pallidum, Right-Thalamus, Right-CaudateRight-Putamen, Right-Pallidum |
| ventricles | 3rd-Ventricle, 4th-Ventricle, Left-Lateral-Ventricle, Left-Inf-Lat-VentRight-Lateral-Ventricle, Right-Inf-Lat-Vent |
| cerebellum | Left-Cerebellum-White-Matter, Left-Cerebellum-Cortex, Right-Cerebellum-White-Matter, Right-Cerebellum-Cortex |
| left_thalamus | Left-Thalamus |
| left_caudate | Left-Caudate |
| left_putamen | Left-Putamen |
| left_globus_pallidus | Left-Pallidum |
| right_thalamus | Right-Thalamus |
| right_caudate | Right-Caudate |
| right_putamen | Right-Putamen |
| right_globus_pallidus | Right-Pallidum |
| icv | CSF, Left-Cerebellum-White-Matter, Left-Cerebellum-Cortex, Right-Cerebellum-White-MatterRight-Cerebellum-Cortex, WM-hypointensities, Left-Cerebral-White-Matter, Left-Cerebral-CortexLeft-Amygdala, Left-Hippocampus, Left-Accumbens-area, Left-HypoThal-noMBLeft-Fornix, Left-MammillaryBody, Left-Basal-Forebrain, Left-SeptalNucRight-Cerebral-White-Matter, Right-Cerebral-Cortex, Right-Hippocampus, Right-AmygdalaRight-Accumbens-area, Right-HypoThal-noMB, Right-Fornix, Right-MammillaryBodyRight-Basal-Forebrain, Right-SeptalNuc, Optic-Chiasm, Left-ThalamusLeft-Caudate, Left-Putamen, Left-Pallidum, Right-ThalamusRight-Caudate, Right-Putamen, Right-Pallidum |

**Supplementary Table 2.** Percentage of segmentations excluded following QC, by template age and cohort. Segmentations were assessed on a region-by-region basis within each subject.

| Region | Template Age |  |  |  |  |
| --- | --- | --- | --- | --- | --- |
| UCT-Khula |  |  |  |  |  |
|  | 3M | 6M | 12M | 18M | 24M |
| Supratentorial tissue | 7.0% | 4.4% | 2.6% | 12.2% | 4.5% |
| Supratentorial CSF | 7.6% | 4.4% | 2.6% | 12.2% | 4.5% |
| Ventricles | 7.6% | 3.9% | 2.6% | 9.8% | 3.4% |
| Cerebellum | 8.3% | 6.9% | 7.2% | 21.9% | 4.5% |
| Cerebellum CSF | 8.3% | 6.9% | 7.8% | 21.9% | 4.5% |
| Brainstem | 8.3% | 7.4% | 7.2% | 21.9% | 4.5% |
| Brainstem CSF | 8.3% | 6.9% | 7.8% | 21.9% | 4.5% |
| Left thalamus | 7.6% | 3.9% | 2.6% | 9.8% | 3.4% |
| Left caudate | 7.6% | 3.9% | 2.6% | 9.8% | 3.4% |
| Left putamen | 7.6% | 3.9% | 2.6% | 9.8% | 3.4% |
| Left globus pallidus | 7.6% | 3.9% | 2.6% | 9.8% | 3.4% |
| Right thalamus | 7.6% | 3.9% | 2.6% | 9.8% | 3.4% |
| Right caudate | 7.6% | 3.9% | 2.6% | 9.8% | 3.4% |
| Right putamen | 7.6% | 3.9% | 2.6% | 9.8% | 3.4% |
| Right globus pallidus | 7.6% | 3.9% | 3.3% | 9.8% | 3.4% |
| Posterior callosum | 8.3% | 3.9% | 3.3% | 9.8% | 3.4% |
| Mid-posterior callosum | 8.3% | 4.4% | 6.5% | 12.2% | 4.5% |
| Central callosum | 7.6% | 3.9% | 4.6% | 12.2% | 4.5% |
| Mid anterior callosum | 7.6% | 4.9% | 2.6% | 14.6% | 4.5% |
| Anterior Callosum | 7.6% | 6.4% | 3.9% | 14.6% | 3.4% |
| Uganda-PRIMES |  |  |  |  |  |
| Supratentorial tissue | 11.5% | 23.5% | 13.4% | - | - |
| Supratentorial CSF | 11.5% | 23.5% | 13.4% | - | - |
| Ventricles | 11.5% | 23.5% | 14.9% | - | - |
| Cerebellum | 11.5% | 23.5% | 13.4% | - | - |
| Cerebellum CSF | 11.5% | 23.5% | 13.4% | - | - |
| Brainstem | 11.5% | 23.5% | 13.4% | - | - |
| Brainstem CSF | 11.5% | 23.5% | 13.4% | - | - |
| Left thalamus | 11.5% | 25.0% | 13.4% | - | - |
| Left caudate | 11.5% | 25.0% | 13.4% | - | - |
| Left putamen | 11.5% | 23.5% | 13.4% | - | - |
| Left globus pallidus | 11.5% | 23.5% | 13.4% | - | - |
| Right thalamus | 11.5% | 25.0% | 13.4% | - | - |
| Right caudate | 11.5% | 26.5% | 13.4% | - | - |
| Right putamen | 11.5% | 23.5% | 13.4% | - | - |
| Right globus pallidus | 11.5% | 23.5% | 13.4% | - | - |
| Posterior callosum | 11.5% | 23.5% | 13.4% | - | - |
| **Mid-posterior callosum** | 11.5% | 23.5% | 13.4% | - | - |
| **Central callosum** | 11.5% | 23.5% | 13.4% | - | - |
| **Mid anterior callosum** | 11.5% | 27.9% | 13.4% | - | - |
| **Anterior Callosum** | 11.5% | 27.9% | 13.4% | - | - |

**Supplementary Table 3.**
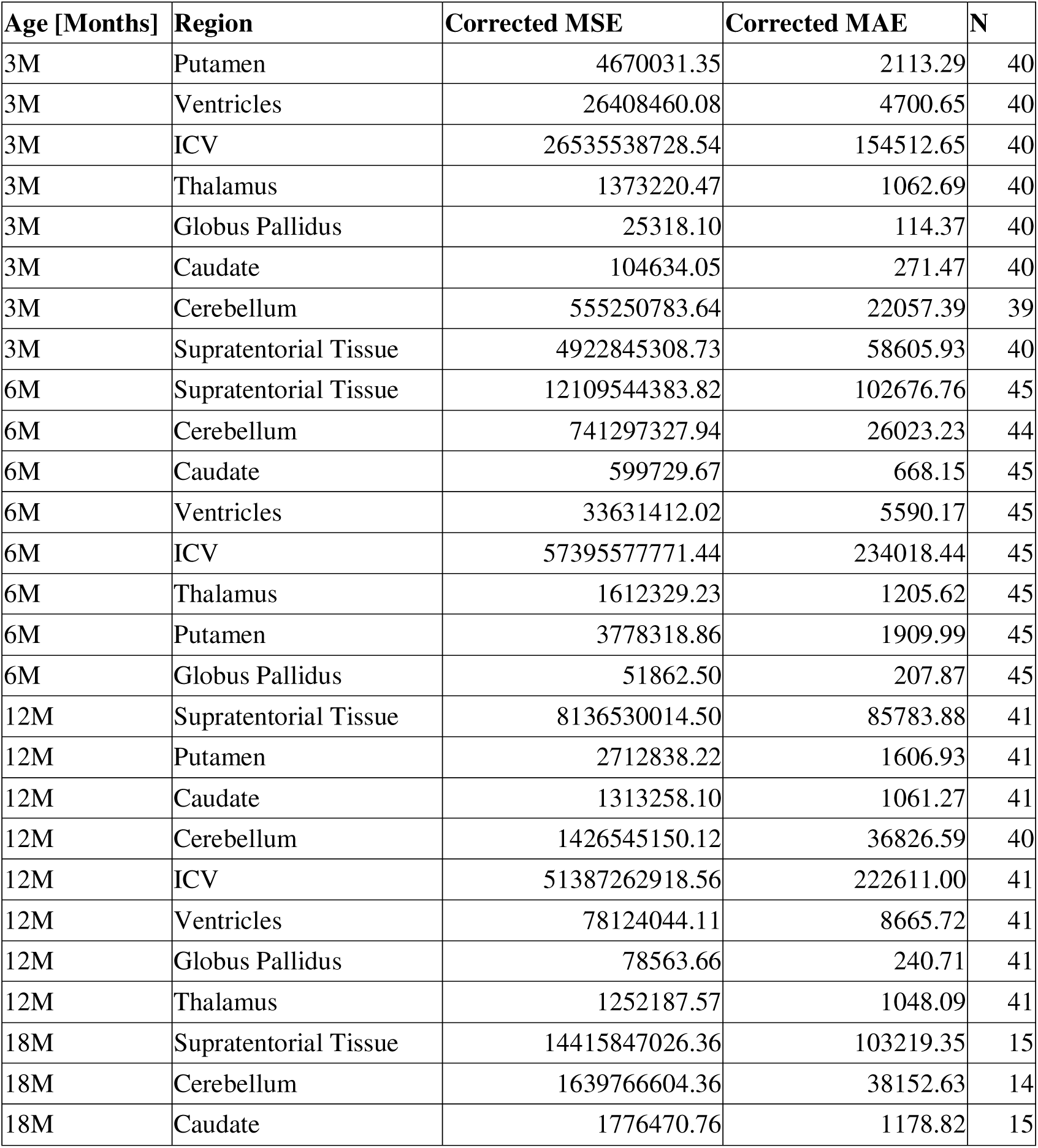
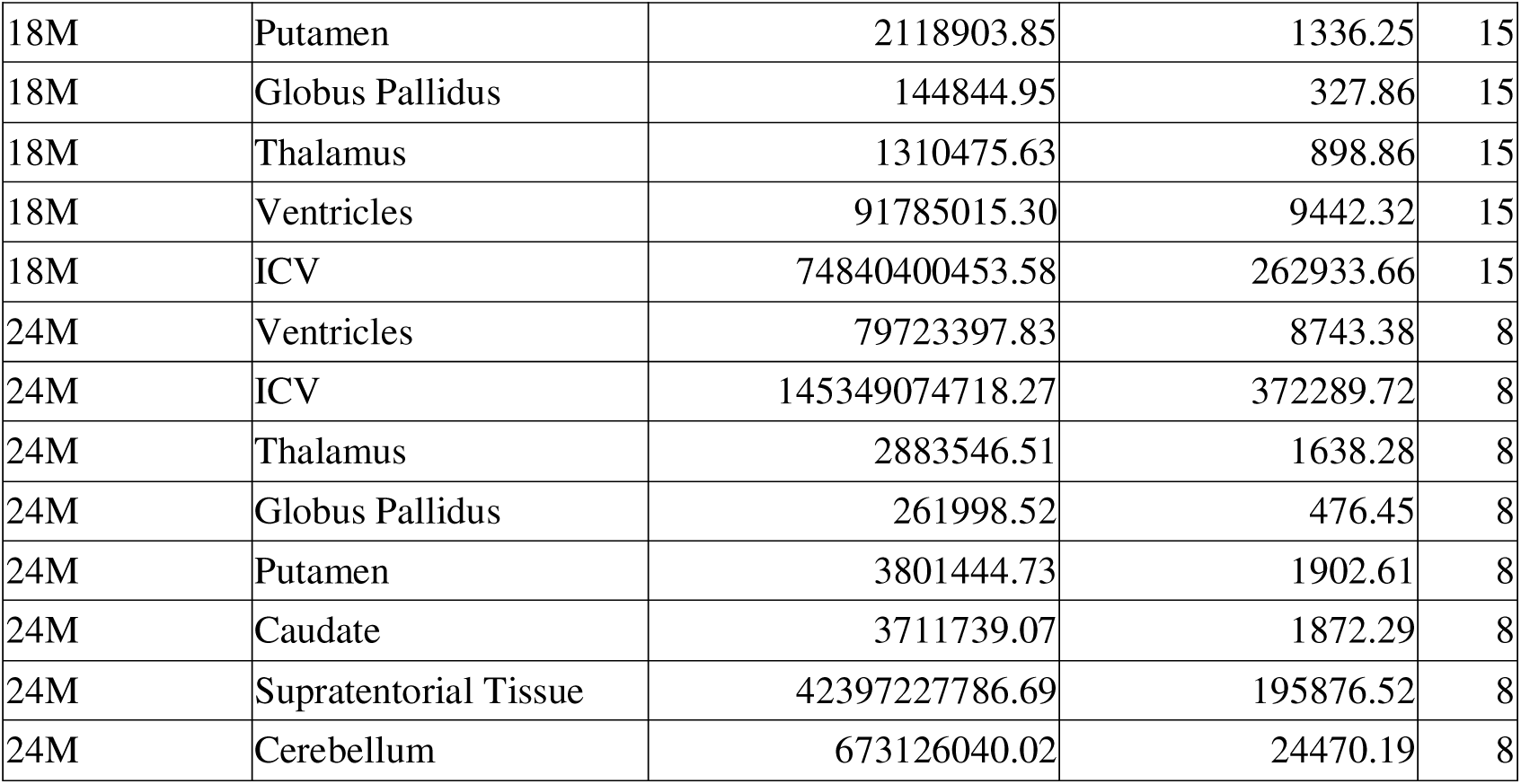
Age corrected mean squared error, age corrected mean absolute error and number of subjects for each region and age between matched scans in the UCT-Khula dataset. Absolute errors were largest for global tissue and head-size measures (e.g., ICV and supratentorial tissue) and for CSF spaces (ventricles), reflecting both their larger anatomical scale and greater sensitivity to boundary/partial-volume effects. In contrast, deep grey nuclei showed substantially smaller errors, with the globus pallidus consistently exhibiting the lowest MAE across ages.

